# WALK for Freezing of Gait in Parkinson’s Disease: A Sham-Controlled Home Rehabilitation Trial

**DOI:** 10.64898/2026.05.14.26352486

**Authors:** Rajesh Benny, Amey Desai, Anil Venkitachalam, Vicky Thakkar, Ria Rajput, Samit Chakrabarty

**Affiliations:** Fortis Hospital Mulund, Mumbai, India; Lifespark Technologies Pvt. Ltd., Mumbai, India; School of Biomedical Sciences, Faculty of Biological Sciences, University of Leeds, United Kingdom

**Keywords:** Parkinson’s disease, freezing of gait, wearable technology, localised muscle-tendon vibration, neuromodulation, home rehabilitation, motor learning, feasibility

## Abstract

**Background:** Freezing of gait (FOG) in Parkinson’s disease often persists despite dopaminergic treatment. WALK is a bilateral thigh-worn device that delivers local muscle-tendon vibration during walking. We examined whether WALK could be incorporated into supervised home rehabilitation and explored differences in freezing severity between active and sham treatment.

**Methods:** In this single-centre pilot randomised clinical trial, adults with Parkinson’s disease and neurologist-confirmed FOG were assigned to Active or Sham-WALK. Both groups completed the same 16-session, six-week supervised home programme; mZ-FOG was assessed in the medication OFF state at baseline and after sessions 8 and 16. At each follow-up, assessment without the device preceded assessment with the assigned device. The complete-case analysis tested the Group × Condition interaction across the five conditions. Trial registration: Clinical Trials Registry-India, CTRI/2022/01/039256, https://ctri.nic.in/; registered 7 January 2022.

**Results:** Ninety people were screened, 47 were randomised (active, n=27; sham, n=20), and 40 completed follow-up and entered the main analysis (active, n=24; sham, n=16). Retention was 85.1% (95% CI 74.9% to 95.4%), and all completers finished the 16-session programme. No serious device-related adverse event was reported among completers; one active-group participant discontinued because of fatigue. mZ-FOG scores decreased in both groups, but the Group × Condition interaction was not significant (F=0.847, p=0.465). At S16 before device reapplication, the baseline-adjusted active-minus-sham difference was −2.98 points (95% CI −6.63 to 0.66; p=0.106); with the assigned device it was −2.87 points (95% CI −6.76 to 1.02; p=0.144). In the Active-WALK group, 18 of 24 improved by at least the 4.28-point MCID.

**Conclusion:** WALK was feasible to integrate into supervised home rehabilitation and was associated with improvement in freezing across training. The lower active-group score before device reapplication at S16 suggests that improvement may have extended beyond the period of immediate stimulation.

## Introduction

Freezing of gait consists of brief episodes in which a person cannot continue moving forwards despite intending to walk. It limits mobility, increases the risk of falling and reduces independence. These effects are not captured well by steady walking in a straight line because freezing commonly occurs during gait initiation, turning, doorway passage, movement through narrow or cluttered spaces, or walking while performing another task [1,2,13–15]. These situations occur repeatedly at home.

Dopaminergic treatment improves some freezing in the medication OFF state, but disabling episodes often remain [2]. Physiotherapy, gait practice, and external cues are therefore important. Their effects vary between people and situations. Cues may also require conscious attention when attention is already divided, for example during turning or moving around obstacles [2,13,14]. A wearable intervention would be most useful if it could accompany practice in these demanding situations without adding much burden.

WALK (Wearable Assisted Locomotor Kinaesthesia) is a bilateral thigh-worn device that applies local mechanical vibration over the muscle-tendon tissue during walking. Unlike visual or auditory cues that must be detected and followed consciously, WALK uses muscle-tendon vibration to recruit proprioceptive afferents from the moving limbs during voluntary walking. This provides sensory input without requiring the participant to follow an additional external timing signal.

The rationale for this approach rests on direct human neurophysiology. Low-amplitude tendon vibration strongly activates primary muscle-spindle endings, often phase-locking their discharge to the vibration cycle, and produces corresponding changes in kinaesthetic perception [31,34]. Vibration also modifies tonic and phasic stretch reflexes, presynaptic control of Ia transmission, agonist-antagonist interactions, and heteronymous spinal reflexes [16,17,20–23,32,33]. These provide a physiological basis for applying vibration during walking, when proprioceptive input and descending locomotor commands converge on the motoneuronal final common pathway [38].

Motoneuronal excitability is also altered in Parkinson’s disease. Alaei and colleagues reported higher initial motor-unit discharge rate and lower recruitment thresholds in people with Parkinson’s disease than in age-matched controls [40]. Measures associated with persistent inward-current amplification were also greater in the medication OFF state and were not fully normalised by dopaminergic medication. These findings show that the motoneuronal state through which sensory and descending inputs are converted into motor output remains altered in Parkinson’s disease.

An earlier prospective, non-randomised study found immediate reductions in freezing severity and task-completion time with WALK, particularly during narrow-space negotiation [27]. That study did not determine whether repeated stimulation could be incorporated into home rehabilitation, whether improvement, carryover effects, would be retained in absence of stimulation, and would the change differ from that associated with rehabilitation. The present pilot addressed these questions by comparing Active and Sham WALK during the same 16-session supervised home programme.

The primary objective here, was to establish whether WALK could be delivered during supervised home rehabilitation, as judged from retention, session completion, home delivery, device tolerance, technical problems and adverse events. The clinical assessments distinguished change during rehabilitation, improvement present without the device at follow-up and additional within-session change after application of the assigned device. The main clinical analysis tested whether mZ-FOG scores changed differently between the active and sham WALK groups across the five assessment conditions. MCID-based categories were used to describe the magnitude and distribution of individual improvement within the active WALK cohort, while task-level scores were used to identify the activities in which changes were most apparent.

## Materials and methods

### Study design and ethics

This was a single-centre, parallel-group, sham-controlled pilot randomised clinical trial. The trial was prospectively registered with the Clinical Trials Registry-India on 7 January 2022 (CTRI/2022/01/039256). The first participant was enrolled on 10 January 2022, and follow-up was completed on 25 October 2025. The Institutional Ethics Committee of Fortis Hospitals Limited, Mumbai, India, approved the study (IEC/2021/OAS/08; 26 November 2021), and all participants provided written informed consent. All supervised training sessions and approximately 90% of assessments took place in participant homes, either by live video or with an investigator present.

### Participants

Adults with a clinical diagnosis of Parkinson’s disease were eligible if they had Hoehn and Yahr stage 2-4 disease, could walk independently and had FOG confirmed by a neurologist. We excluded people whose walking difficulty was mainly caused by another neurological or orthopaedic condition, whose medical condition was not adequately controlled, or whose medical, psychiatric, cognitive or communication needs made safe participation or adherence unlikely. People were also excluded if they could not perform the study tasks safely or could not wear the device. Participants continued their usual clinical care. Assessments were performed in the medication OFF state, defined in the protocol as at least 4 h after the last dopaminergic dose. In practice, assessment was usually scheduled 15–30 min before the next dose or after omission of a scheduled dose, provided that the 4 h interval had been met.

### Randomisation, concealment, and masking

A computer-generated sequence, stratified by Hoehn and Yahr stage, used a planned 1:1 allocation to Active or Sham-WALK. The realised allocation was 27 participants to Active-WALK and 20 to Sham-WALK. An independent coordinator who was not involved in recruitment or assessment held the allocation sequence and communicated the assigned device mode to the intervention team. Participants were told that two wearable conditions were being compared but were not told their allocation. Outcome assessors were not informed of the allocation or device mode during live scoring. Therapists could not be masked because vibration was perceptible.

### WALK intervention

WALK comprised two thigh-worn units that applied local mechanical vibration over the underlying muscle-tendon tissues. During set-up, each participant completed a one-minute walking trial. A clinician measured cadence from this trial and used the measurement to configure the device, either at the study site or remotely when needed. Cadence was used only for initial configuration during the first session and was not measured continuously or updated automatically during treatment in this study.

Participants in both groups wore physically identical units. In the Active-WALK group, local muscle-tendon vibration was enabled during walking; in the Sham-WALK group, the same units were worn with vibration disabled. WALK was intended to alter proprioceptive input during voluntary walking rather than to generate a step or evoke muscle contraction directly. Figure 1 shows the device and the proposed physiological mechanism of action. Firmware, controller settings, and other commercially sensitive technical specifications are proprietary.

**Figure 1.**
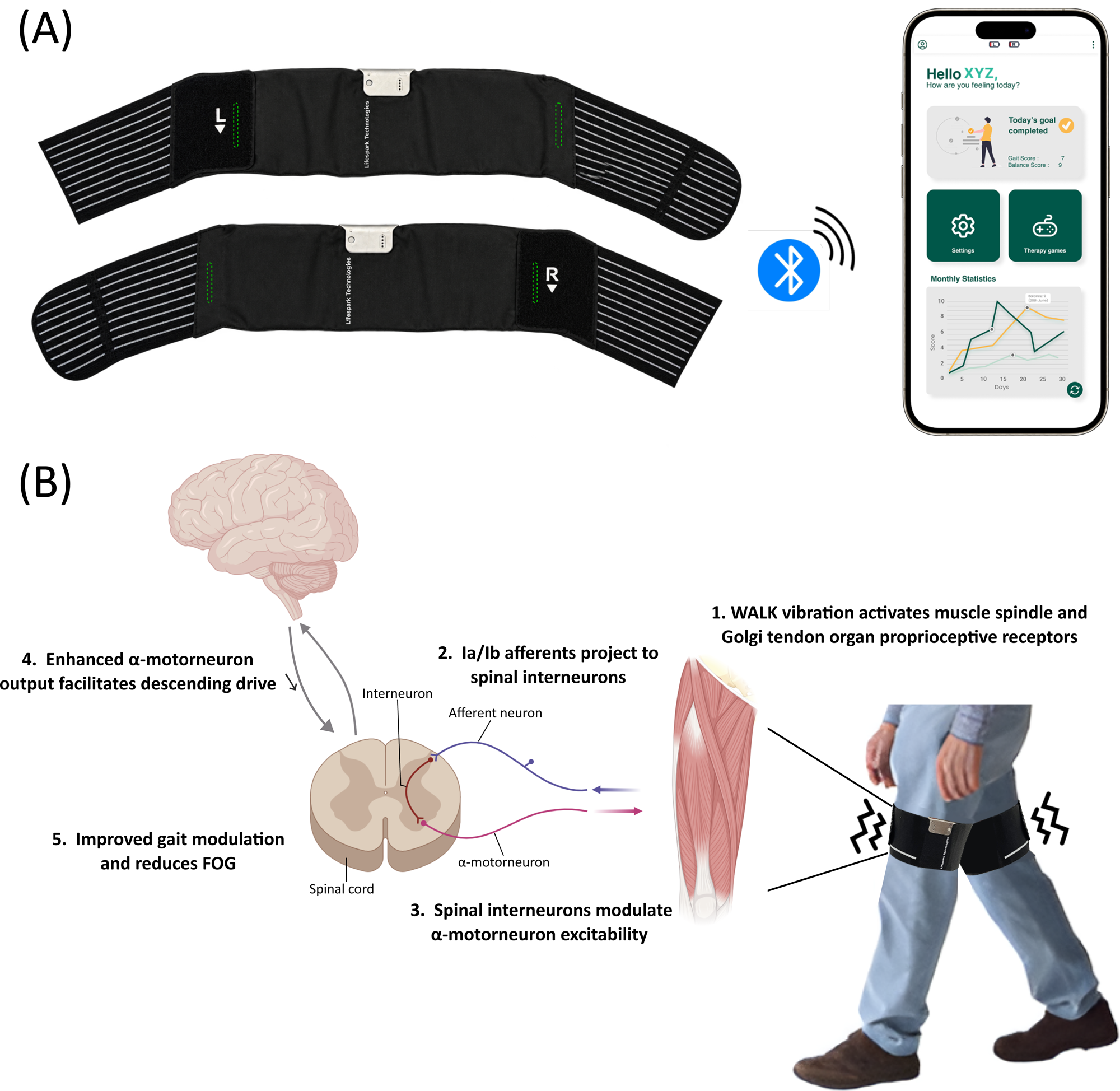
WALK system and physiological basis of the intervention. (A) Bilateral thigh-worn WALK units and wireless configuration interface. (B) Local muscle-tendon vibration recruits proprioceptive afferents, which project to spinal sensorimotor circuits where sensory and descending locomotor inputs converge on motoneurons. The proposed downstream effects include altered sensorimotor processing, more effective expression of residual descending locomotor drive, improved step initiation and reduced freezing. Proprioceptive-afferent recruitment by muscle-tendon vibration is well established; the downstream pathway proposed for WALK was not measured directly in this trial.

**Figure 2.**
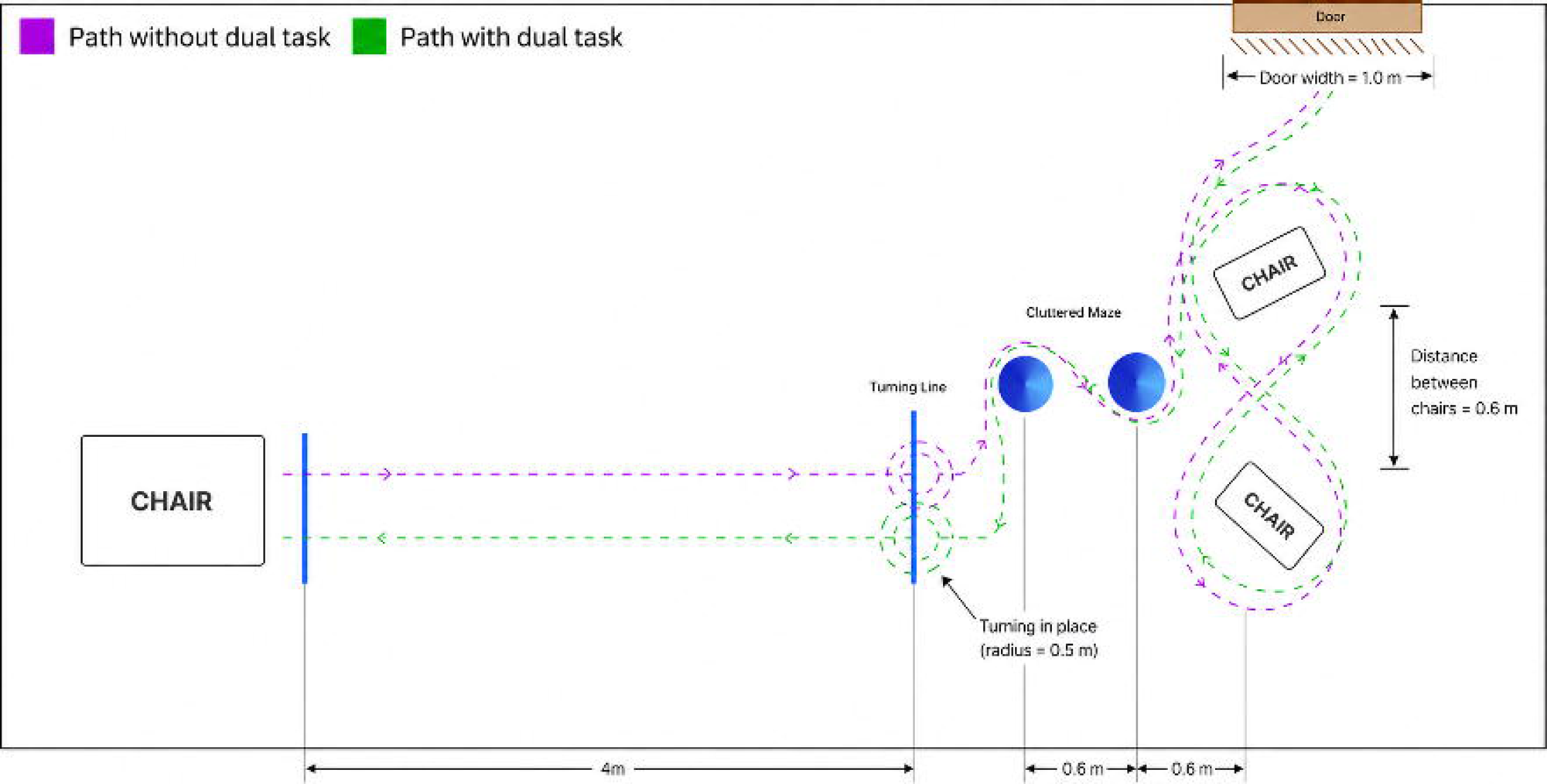
Modified Ziegler-based FOG assessment course. The course included 4 m of straight walking, in-place turns, negotiation of a cluttered maze and passage through a 1.0 m doorway under single-task and cognitive dual-task conditions.

### Home-based motor-learning programme

Both groups completed the same 16-session home-based programme spread over six weeks. Sessions lasted approximately 40–50 minutes and were supervised throughout. Assessments were conducted after eight completed sessions and after completion of all sixteen sessions. Training included weight shifting, starting and stopping, straight walking, turning, figure-of-eight walking, passing through doorways, negotiating cluttered paths and walking while performing a cognitive task. The supervising therapist progressed the exercises according to each participant’s ability to complete them safely by increasing task complexity within the specified activities. Both groups received the same programme and task progression. The only planned difference was device mode: vibration was enabled during training in the Active-WALK group and disabled in the Sham-WALK group.

### Assessment schedule and outcomes

All participants were assessed without the device at baseline. At S8 and S16, each participant completed two assessments in a fixed order, first without the device and then with the assigned active or sham device. This fixed order was chosen because preliminary use suggested that active stimulation might influence a later assessment if the device condition came first. The order reduced the risk of carryover into the without-device assessment, but it also meant that practice, fatigue, and test order could affect the comparison made within the same session.

The main clinical outcome was freezing severity measured with a modified Ziegler-based task battery (mZ-FOG). The Ziegler test is a validated clinician-rated measure of FOG [3,5], and the original 0-3 item-level scoring system was retained for each item. Participants completed six tasks, under single-task and cognitive dual-task conditions: standing up and sitting down, walking 4 m in a straight line, completing 360° turns, 540° turns, negotiating a cluttered maze, and passing through a doorway. The 12 item scores were summed to give a total from 0 to 36 with higher scores indicating more severe freezing.

The cluttered maze created repeated changes in available walking space and direction, bringing together narrow-space negotiation, turning and obstacle avoidance within one task. These are recognised FOG-provoking circumstances, and expert consensus identifies turning hesitation, narrow-space hesitation, start hesitation and cognitive dual-tasking as core conditions for clinician-rated assessment of FOG severity [39]. The expanded battery was therefore designed to sample situations that commonly provoke FOG at home.

Secondary clinical outcomes assessed balance, gait, mood, anxiety and autonomic symptoms using the Mini-BESTest [6–8], Functional Gait Assessment [9,10], Tinetti Performance-Oriented Mobility Assessment [11], Hamilton Depression Rating Scale, Geriatric Anxiety Scale and COMPASS-31 [12]. Feasibility outcomes included recruitment, retention, completion of assessments and supervised sessions, delivery of the home-based programme, device tolerance, technical problems, protocol deviations, and adverse events.

Individual improvements within the Active-WALK group were evaluated using the MCID of 4.28 mZ-FOG points reported in the earlier WALK study [27]. Participants were grouped by the magnitude of improvement: more than twice the MCID (>8.56 points), one to two times the MCID (4.28–8.56 points), less than one MCID (>0 to <4.28 points), or no improvement or worsening (≤0 points). Analyses using the exact 4.28-point value and the rounded 4-point equivalent produced the same participant classification. The analysis was designed to characterise the magnitude and distribution of individual improvement within the Active WALK cohort.

### Outcome scoring and safety

Assessments were video recorded for documentation and later review but were scored live during the home sessions. During each assessment, investigators monitored falls, near-falls, skin irritation, device-related discomfort, dizziness, fatigue, and device malfunction. Participants could pause or stop at any time. If a task was stopped for safety, the score recorded up to that point was kept; outcomes that could not be observed were recorded as missing.

### Sample size and statistical analysis

This pilot study assessed feasibility and estimated the size and variability of clinical change to inform a larger trial. Analyses were performed using R version 4.5.3 and JASP version 0.19.3. The main analysis included the 40 participants with mZ-FOG scores available in all five assessment conditions: baseline without the device, S8 without and with the assigned device, and S16 without and with the assigned device. Participants were analysed in their randomised groups.

A mixed repeated-measures ANOVA tested whether change across the five conditions differed between the Active and Sham WALK groups. Sphericity was assessed using Mauchly’s test, with the Greenhouse–Geisser correction applied when required. Linear mixed-effects, rank-normal, bootstrap and baseline-adjusted analyses assessed the consistency of the findings across statistical methods. Condition-specific estimates, within-session comparisons and task-level analyses were exploratory, with Bonferroni adjustment applied to the specified pairwise comparisons. The MCID-based analysis described individual improvement within the Active-WALK cohort. Missing values were not imputed, and each analysis used the available data for the relevant outcome.

## Results

### Participants and baseline characteristics

Ninety people were screened and 47 were randomised, with 27 assigned to Active-WALK and 20 to Sham-WALK (Figure 3). Forty participants completed follow-up and mZ-FOG scores were obtained in all five assessment conditions for 24 participants in the Active-WALK group and 16 in the Sham-WALK group. These 40 participants were included in the main repeated-measures analysis and analysed according to their randomised group. Seven participants did not complete the scheduled follow-up assessments. In the Active-WALK group, one was lost to follow-up, one had no available visit data and one discontinued because of fatigue. In the Sham-WALK group, two were lost to follow-up, one had no available visit data, and one withdrew because no effect was perceived.

**Figure 3.**
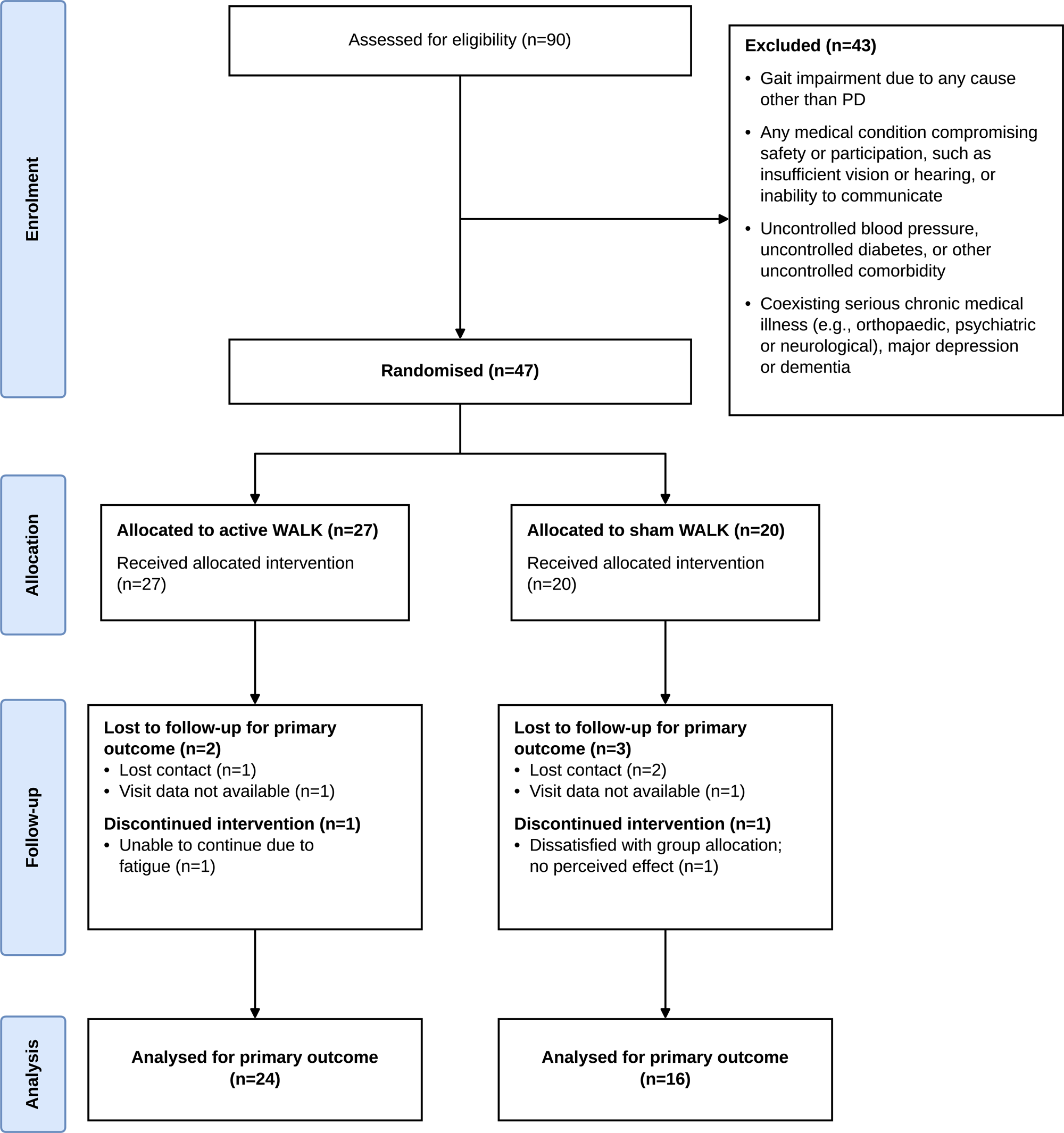
Participant flow and analysis population. Ninety people were screened and 47 were randomised: 27 to active WALK and 20 to sham WALK. Forty participants completed all five mZ-FOG assessments and entered the complete-case repeated-measures analysis in their randomised groups: active, n=24; sham, n=16. Seven randomised participants did not complete follow-up; reasons are shown by group.

Baseline mZ-FOG scores were similar in the active and Sham-WALK groups at 16.42 ± 7.64 and 16.63 ± 8.37, respectively. Age, disease duration and Hoehn and Yahr stage were also similar, although the Active-WALK group included a higher proportion of men (Table 1).

**Table 1.** Baseline demographic and clinical characteristics of participants included in the main analysis.

| Characteristic | Active-WALK (n=24) | Sham-WALK (n=16) |
| --- | --- | --- |
| Age, years | 68.0 ± 6.5 | 66.3 ± 8.8 |
| Sex, n (%) | Male 22 (91.7%); Female 2 (8.3%) | Male 11 (68.8%); Female 5 (31.2%) |
| Disease duration, years | 10.3 ± 7.5 | 9.8 ± 5.1 |
| Hoehn and Yahr stage, median [IQR] | 3 [2.9, 3.0] | 3 [3.0, 3.0] |
| mZ-FOG total, 0–36 | 16.4 ± 7.6 | 16.6 ± 8.4 |
| Tinetti/POMA, 0–28 | 20.1 ± 3.7 | 19.6 ± 5.0 |
| Mini-BESTest, 0–28 | 18.6 ± 3.9 | 17.0 ± 5.3 |
| Functional Gait Assessment, 0–30 | 15.3 ± 5.7 | 14.6 ± 6.1 |
| Hamilton Depression Rating Scale | 7.4 ± 4.5 | 8.9 ± 6.7 |
| COMPASS – 31, 0–100 | 12.9 ± 7.7 | 8.8 ± 6.4 |
| Geriatric Anxiety Scale | 17.0 ± 9.9 | 17.3 ± 14.7 |

| Characteristic | Active-WALK (n=24) | Sham-WALK (n=16) |
| --- | --- | --- |
| Age, years | 68.0 ± 6.5 | 66.3 ± 8.8 |
| Sex, n (%) | Male 22 (91.7%);<br>Female 2 (8.3%) | Male 11 (68.8%);<br>Female 5 (31.2%) |
| Disease duration, years | 10.3 ± 7.5 | 9.8 ± 5.1 |
| Hoehn & Yahr stage,<br>median [IQR] | 3 [2.9, 3.0] | 3 [3.0, 3.0] |
| mZ-FOG total (S0), 0–36 | 16.4 ± 7.6 | 16.6 ± 8.4 |
| Tinetti / POMA, 0–28 | 20.1 ± 3.7 | 19.6 ± 5.0 |
| Mini-BESTest, 0–28 | 18.6 ± 3.9 | 17 ± 5.3 |
| Functional Gait Assessment<br>(FGA), 0–30 | 15.3 ± 5.7 | 14.6 ± 6.1 |
| Hamilton Depression Rating<br>Scale (HDRS) | 7.4 ± 4.5 | 8.9 ± 6.7 |
| COMPASS-31, 0–100 | 12.9 ± 7.7 | 8.8 ± 6.4 |
| Geriatric Anxiety Scale<br>(GAS) | 17.0 ± 9.9 | 17.3 ± 14.7 |

Values are mean ± SD unless otherwise stated. Hoehn and Yahr stage is reported as median [IQR]. Higher mZ-FOG scores indicate more severe freezing. Higher Mini-BESTest, Functional Gait Assessment and Tinetti/POMA scores indicate better performance; lower HDRS, GAS and COMPASS-31 scores indicate fewer symptoms. Baseline assessments were conducted in the medication OFF state

### Feasibility and safety

Forty of the 47 randomised participants completed follow-up, giving a retention rate of 85.1% (95% CI 74.9% to 95.4%). All 40 completers finished the 16-session programme. All training sessions and approximately 90% of assessments were conducted in participant homes.

No participant completing follow-up discontinued because of device intolerance, and no serious device-related adverse event was reported among the 40 completers (0/40; exact 95% CI 0% to 8.8%). No falls occurred during supervised assessments and skin checks were unremarkable. One participant in the Active-WALK group discontinued owing to fatigue and was excluded from the completer analysis. The retention rate, completion of all sessions by follow-up completers and low incidence of device-related problems support the feasibility of integrating WALK into supervised home rehabilitation.

### Primary clinical outcome

mZ-FOG scores decreased in both groups across the five assessment conditions (Figure 4). There was a significant effect of Condition (F=15.94, p<0.001, partial η^2^=0.295), indicating that mZ-FOG scores changed across the five assessment conditions. The Group × Condition interaction was not significant (Greenhouse-Geisser-corrected F=0.847, p=0.465, partial η^2^=0.022), and there was no overall Group effect (F=0.904, p=0.348, partial η^2^=0.023). Linear mixed-effects and other sensitivity analyses produced the same overall interpretation.

**Figure 4.**
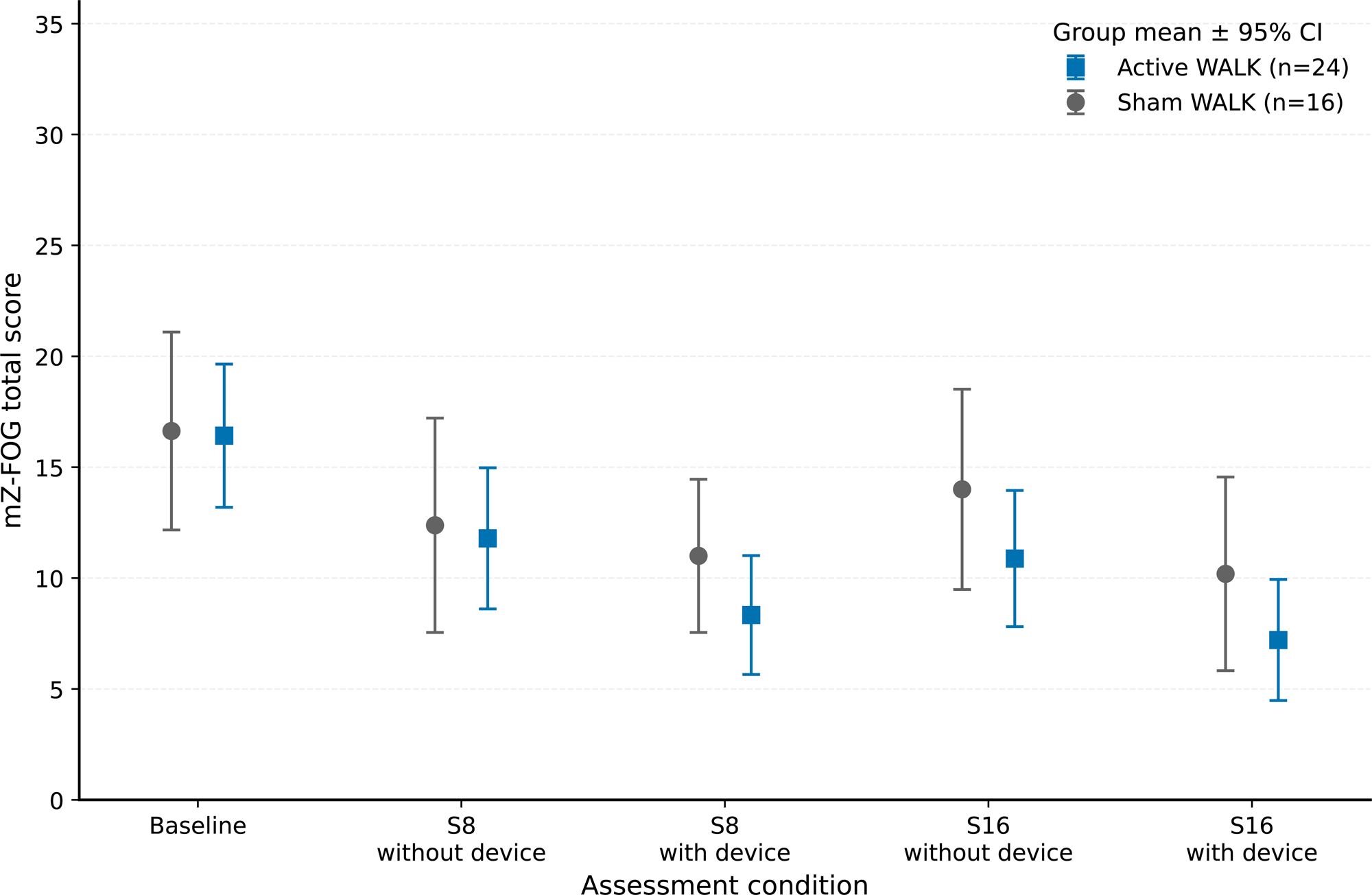
Total mZ-FOG scores across the five assessment conditions. Points show group means and error bars show 95% confidence intervals for the 40 participants with complete data (active, n=24; sham, n=16). Lower scores indicate less severe freezing. The Group × Condition interaction across all five conditions was not significant (p=0.465). At S16 before device reapplication, the baseline-adjusted active-minus-sham difference was −2.98 points (95% CI −6.63 to 0.66; p=0.106); with the assigned device, it remained similar at −2.87 points (95% CI −6.76 to 1.02; p=0.144). The S16 difference was therefore already present before immediate stimulation at the final assessment.

### Between-group estimates

The assessment-specific Active-minus-Sham estimates were small at S8 without the device and greater at the subsequent conditions. At S16 before the device was reapplied, the baseline-adjusted active-minus-sham difference was −2.98 mZ-FOG points (95% CI −6.63 to 0.66; p=0.106; standardised difference −0.535). The Active-WALK group therefore had lower freezing scores than the Sham-WALK group before immediate device use at the final assessment.

With the assigned device, the adjusted S16 difference remained similar at −2.87 points (95% CI −6.76 to 1.02; p=0.144; standardised difference −0.482). Earlier adjusted differences were −0.46 points at S8 without the device and −2.57 points at S8 with the assigned device (Supplementary Table S4).

The confidence intervals included zero, so the size of the between-group difference remains uncertain. The similar S16 estimates before and after device reapplication indicate that the observed separation was already present before immediate stimulation at the final assessment.

### Change over time by group

The mZ-FOG scores decreased in both groups, although the pattern differed across assessments (Figure 4). In the Active-WALK group, the mean score decreased from 16.42 ± 7.64 at baseline to 11.79 ± 7.54 before device use and 8.33 ± 6.35 with the assigned device at S8. At S16, the corresponding scores were 10.88 ± 7.27 and 7.21 ± 6.47. The S16 score obtained before the device was reapplied was 5.54 points below baseline (95% CI 1.21 to 9.88; p=0.003).

In the Sham-WALK group, the mean score decreased from 16.63 ± 8.37 at baseline to 12.38 ± 9.07 before device use and 11.00 ± 6.48 with the assigned device at S8. At S16, the corresponding scores were 14.00 ± 8.48 and 10.19 ± 8.19.

At S8, the within-session reduction was 3.46 points with Active-WALK and 1.38 points with Sham-WALK. At S16, the corresponding reductions were similar at 3.67 and 3.81 points. By S16, however, the Active-WALK group already had lower scores before the device was reapplied. This pattern suggests an early immediate reduction in freezing to Active-WALK followed by a later improvement that was present before stimulation.

### Improvement within the active WALK cohort

Among the 24 Active-WALK completers, 18 participants (75%) improved by at least the 4.28-point MCID from baseline with the assigned device. Fourteen participants (58.3%) improved by more than twice the MCID, four (16.7%) improved by one to two MCIDs, three (12.5%) improved by less than one MCID, and three (12.5%) showed no improvement or worsened (Supplementary Figure S1).

### Task-specific and secondary outcomes

Greater reductions in freezing were observed under dual-task conditions in the active WALK group across several activities (Figure 5). The pattern was therefore not confined to one walking task. Secondary clinical outcomes are summarised in Supplementary Table S5.

**Figure 5.**
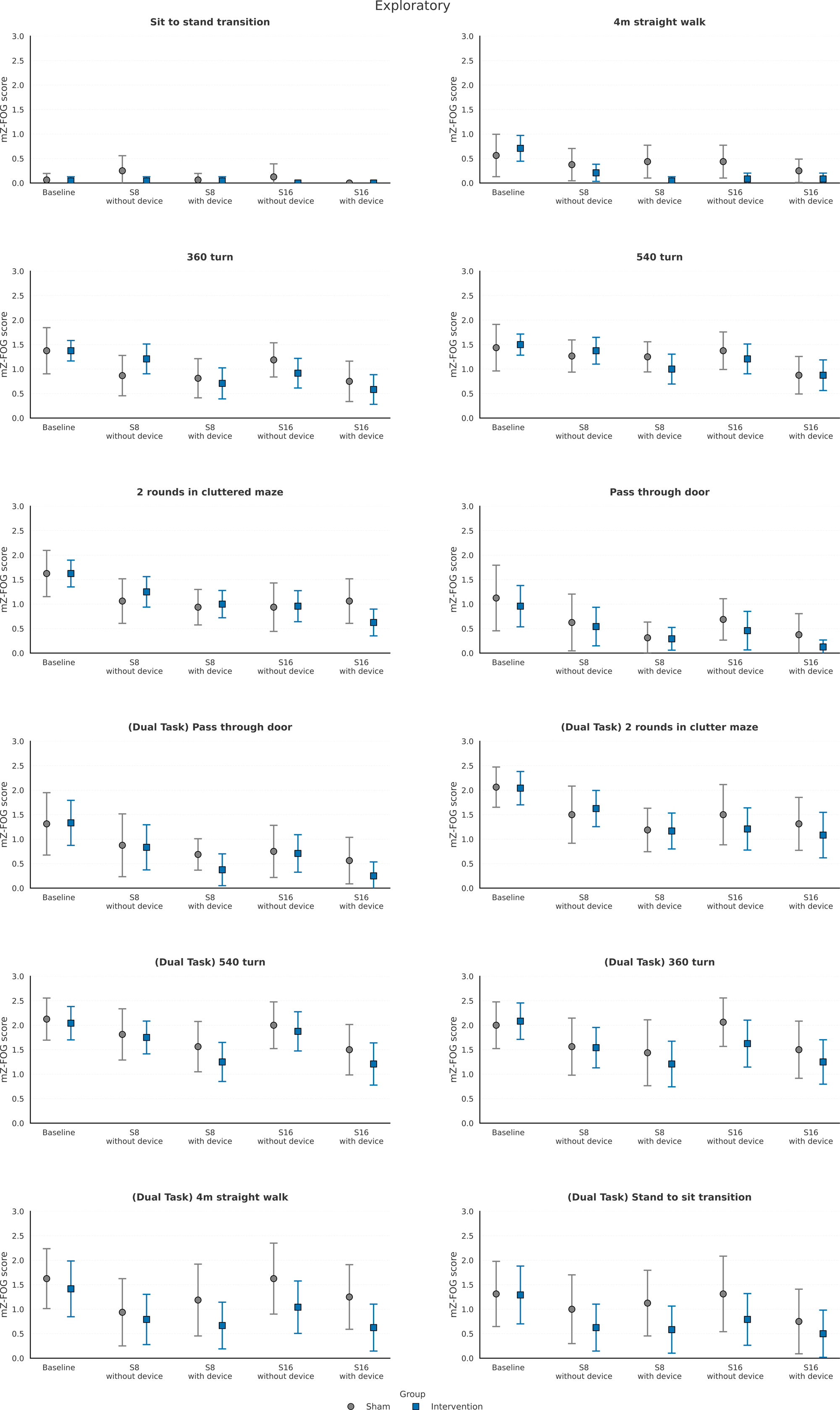
Exploratory mZ-FOG scores for individual walking tasks under single-task and cognitive dual-task conditions. Active and sham WALK groups are plotted on the same 0–3 item-score scale. The profiles show where changes occurred and identify tasks for evaluation in a larger trial; they do not establish a task-specific treatment effect.

## Discussion

### Improvement with WALK changed over training

The improvement associated with active WALK changed over the course of training (Figure 4). At S8, active WALK produced a greater within-session reduction in freezing than sham WALK. By S16, the Active-WALK group already had a lower freezing score before device reapplication, while the immediate reductions with active and sham WALK were similar. The comparable between-group estimates before and during device use at S16 suggest that improvement may have extended beyond the period of immediate stimulation.

Both groups improved during the programme, indicating an important contribution from repeated task practice and supervised rehabilitation. WALK was successfully integrated into home rehabilitation, with 85.1% retention, completion of all 16 sessions by follow-up completers and few device-related problems. At S16, the Active-WALK group had a lower freezing score before the device was reapplied. This timing is important because it suggests that improvement associated with repeated training may have remained present when immediate stimulation was absent.

### Home-based rehabilitation contributed to the improvements

The home setting allowed participants to practise walking situations that commonly provoke FOG under therapist supervision. Task difficulty was adjusted to each participant’s ability, and safety was monitored throughout. Completion of all planned sessions indicated that the programme could be delivered successfully at home.

mZ-FOG scores decreased in both groups (Figure 4), indicating that repeated task practice and therapist supervision contributed to improvement. Active-WALK was therefore evaluated as an addition to the same rehabilitation programme rather than against no treatment.

At S8, the greater within-session reduction with active WALK was consistent with the acute effect of vibration reported previously [27]. At S16, the within-session reductions with active and sham WALK were similar, while the active group had a lower score before device reapplication. The findings therefore suggest a change in the temporal pattern of improvement across training. Follow-up after the programme will establish how long any improvement remains

### Greater improvement in performance across dual-task conditions

Freezing decreased more under dual-task conditions across several activities in the active WALK group (Figure 5). Because the pattern was not restricted to one task, it may reflect improvement in motor-control processes shared across the task battery, including step timing, coordination, adjustment of movement direction, and adaptation to changing spatial demands when attention is divided.

A larger trial should determine whether these improvements are reproduced across participants and activities. If confirmed, the findings would suggest that WALK supports motor control under divided attention rather than improving performance only on a specific trained task. This would strengthen the potential value of WALK for everyday mobility.

### Physiological basis for improvement with WALK

The present findings suggest that repeated vibration delivered during training may have effects beyond the immediate stimulation period. Active WALK produced a greater within-session reduction in freezing at S8, whereas by S16 the difference between groups was already present before device reapplication (Figure 4).

Pereira and colleagues reported that muscle vibration can reduce freezing under specific experimental conditions [29]. During constrained stepping, freezing duration decreased when vibration was applied to the triceps-surae of the less affected limb after freezing had begun. The study demonstrated that altering proprioceptive input can influence freezing behaviour, although it did not identify the neural pathway responsible.

WALK extends this observation from an acute experimental manipulation to repeated wearable delivery. It applies bilateral thigh-based vibration during voluntary movement and throughout rehabilitation, rather than applying vibration reactively to a single distal muscle after freezing begins. The present findings suggest that repeated use may produce an improvement that outlasts immediate stimulation, as lower freezing scores were already present before the device was reapplied at S16.

It has long been established that tendon vibration recruits primary muscle-spindle afferents and alters kinaesthetic perception [31,34]. Tendon vibration also modifies stretch-reflex transmission, presynaptic regulation of Ia afferent input, agonist-antagonist interactions, and heteronymous spinal reflex interactions [16,17,20–23,32,33]. Repeated focal vibration can produce longer-lasting changes in specific spinal circuits [35], while sensory and corticospinal inputs can modify proprioceptive transmission to motoneurons [36,37]. Together, this evidence shows that focal vibration can modulate sensorimotor processing within spinal circuits, where sensory and descending inputs converge on the motoneuronal final common pathway [38].

More direct evidence in Parkinson’s disease comes from Silva-Batista and colleagues, who found that vibration increased presynaptic inhibition of the soleus pathway during step initiation in people with Parkinson’s disease and FOG [30]. This change was associated with a smaller anticipatory postural adjustment. Although the stimulation site and protocol differed from WALK, the study was consistent with the concept that vibration can modify spinal sensorimotor control during preparation for movement.

Recent motor-unit evidence further shows that the spinal motor system is altered in Parkinson’s disease. Alaei and colleagues found higher initial motor-unit discharge rates and lower recruitment thresholds in people with Parkinson’s disease than in age-matched controls [40]. Measures associated with persistent inward-current amplification were also greater in the medication OFF state and were not fully normalised by dopaminergic medication. Although these recordings were obtained from an upper-limb muscle during controlled isometric contractions, the findings show that motoneuronal recruitment and discharge behaviour remain altered in Parkinson’s disease across medication states.

This physiological evidence provides a plausible framework for the clinical pattern seen with WALK. Repeated vibration-driven proprioceptive input may modify how spinal sensorimotor circuits integrate sensory information with descending locomotor commands. Over the course of training, this may improve the timing, coordination, and adjustment of stepping, particularly when attention is divided. Such a mechanism could explain why improvement was initially greater during active stimulation at S8 but was already present before stimulation at S16. Direct recordings of lower-limb motor-unit activity and spinal reflex transmission during WALK use are needed to verify this proposed mechanism directly.

### Strengths and limitations

The study tested WALK during supervised rehabilitation in participant homes. Both groups used physically identical hardware, completed the same rehabilitation programme, and were assessed in the medication OFF state. Allocation was concealed, and most training and assessment took place at home. Assessing participants without the device before testing them with the assigned device allowed improvement already to present at follow-up to be distinguished from change during immediate device use.

The main analysis included the 40 participants with complete mZ-FOG data across all five assessment conditions. The sample was small, and the realised group sizes were unequal. Although the devices appeared identical, the sham condition did not reproduce the sensation of vibration. Therapists could not be masked, and the success of participant masking was not assessed. Assessments followed a fixed order and were scored live.

The mZ-FOG battery extended the validated Ziegler framework to capture a broader range of FOG-provoking activities. The MCID of 4.28 mZ-FOG points reported previously [27] provided a consistent basis for describing individual improvement within the Active-WALK cohort. The responder analysis was designed to characterise the magnitude and distribution of change within that cohort.

At S16, the Active-WALK group had lower freezing scores than the Sham-WALK group before the device was reapplied, suggesting that the observed improvement may have extended beyond the period of immediate stimulation. The study did not establish how long improvement persisted after the rehabilitation programme ended. Because all rehabilitation sessions were supervised, the feasibility findings apply to supervised home delivery rather than independent use of WALK

## Conclusion

WALK was integrated successfully in a supervised 16 session home-rehabilitation programme, with 85.1% retention, completion of all sessions by follow-up completers, and few device-related problems. Active-WALK produced a larger within-session reduction in freezing at S8. By S16, the active group had a lower score before device reapplication, and the between-group estimate remained similar during device use. This pattern suggests that improvement may have extended beyond the period of immediate stimulation.

Within the Active-WALK group, 18 of 24 participants improved by at least the 4.28-point MCID, including 14 who improved by more than twice this threshold. Greater reductions in freezing were also observed across dual-task activities. These findings suggest that repeated rehabilitation with Active-WALK may produce improvement beyond immediate stimulation and may support motor-control processes shared across several challenging activities.

Vibration-driven proprioceptive input provides a plausible physiological basis for these effects through modulation of the sensorimotor processing within spinal circuits. Repeated vibration may allow residual descending locomotor commands to produce more effective motor output. Further evaluation should establish the consistency and duration of improvement and test the proposed mechanism directly.

### Implications for a larger trial

A larger trial should assess participants without the device at the end of training and again after supervised rehabilitation has stopped. This would determine whether the improvement observed at S16 persists and for how long. The home-based assessment framework should be retained, with the 4.28-point MCID applied to both active and Sham-WALK groups. Reporting mean between-group changes alongside the distribution of individual improvement would establish the size and consistency of benefit across a larger population.

The greater reductions in freezing under dual-task conditions in the Active-WALK group should also be examined across a larger population (Figure 5). A sham condition that reproduces vibration sensation, formal assessment of masking and central blinded video scoring would help distinguish the contribution of active vibration from practice and expectation. A mechanistic sub-study should examine lower-limb motor-unit recruitment and discharge behaviour together with conditioned H-reflexes and other measures of spinal gating.

## Supporting information

participants data

consort checklist

TIDier Checklist

## Data Availability

All data produced in the present study are available upon reasonable request to the authors.

## Declarations

### Ethics statement

The study was approved by the Institutional Ethics Committee of Fortis Hospitals Limited, Mumbai, India (IEC/2021/OAS/08). All participants provided written informed consent.

### Author contributions

Conceptualisation: AD, SC. Investigation: RB, AV, VT. Data curation and formal analysis: RR, AD, SC. Methodology: AD, SC. Supervision: SC. Writing, original draft: AD, RR, SC. Writing, review and editing: all authors.

### Data availability statement

De-identified participant data supporting the main findings, together with the data dictionary and analysis code, may be requested from the corresponding author. Access will depend on ethics and institutional approval, the limits of participant consent, an appropriate data-use agreement and protection of commercially confidential intellectual property. Proprietary device firmware, source code, controller settings and other commercially sensitive details are not publicly available. The Methods and Figure 1 provide the information needed to understand how WALK was used in the trial.

### Generative AI statement

Microsoft 365 Copilot (enterprise version) was used for language editing, clarity, and consistency of the manuscript. It was not used to generate or analyse study data, conduct statistical analyses or determine scientific interpretation. All AI-assisted content was reviewed and edited by the authors, who take full responsibility for the final manuscript.

---

**Supplementary Figure S1. MCID-based improvement within the Active-WALK cohort.** Improvement in mZ-FOG score was calculated from baseline without the device to S16 with active WALK for the 24 participants with complete data at both assessments. Categories were defined using the 4.28-point MCID reported in the earlier WALK study [27]: improvement >2×MCID (>8.56 points), improvement 1–2×MCID (4.28–8.56 points), improvement <1×MCID (>0 to <4.28 points), and no improvement or worsening (≤0 points). Fourteen participants (58.3%) improved by >2×MCID, four (16.7%) improved by 1–2×MCID, three (12.5%) improved by <1×MCID, and three (12.5%) showed no improvement or worsened. The analysis was designed to characterise the magnitude and distribution of individual improvement within the active WALK cohort.

**Supplementary Table S1**. Repeated-measures analysis of total mZ-FOG scores across the five assessment conditions.

The analysis included the 40 participants with complete mZ-FOG scores across all fiveons: Active-WALK, n=24; Sham-WALK, n=16. The Group × Condition interaction was the main comparison. Greenhouse–Geisser correction was applied where the assumption of sphericity was not met. Partial eta-squared is reported as the measure of effect size.

**Supplementary Table S2. Linear mixed-effects analysis of total mZ-FOG scores across assessment conditions.**

**Supplementary Table S3.** Bootstrap and rank-normal analyses of changes in total mZ-FOG score.

Within-session contrasts compare scores obtained with the assigned device against scores obtained without the device at the same assessment. Negative values indicate lower mZ-FOG scores with the assigned device. Between-group contrasts are reported as Active-WALK minus Sham-WALK, with negative values favouring Active-WALK. Bootstrap confidence intervals were calculated using 2,000 resamples.

**Supplementary Table S4.**Baseline-adjusted active-versus-sham comparisons at S8 and S16.

Values are estimated means adjusted for baseline mZ-FOG score. Between-group differences are calculated as Active-WALK minus Sham-WALK. Negative values favour Active-WALK because lower mZ-FOG scores indicate less severe freezing. Standardised differences are reported with negative values favouring Active-WALK.

**Supplementary Table S5**

Supplementary Table S5. Secondary clinical outcomes at baseline, S8 and S16.

Values are reported by randomised group and assessment. The direction of improvement differs between measures. Higher scores indicate better performance for the Mini-BESTest, Functional Gait Assessment and Tinetti Performance-Oriented Mobility Assessment. Lower scores indicate fewer symptoms for the Hamilton Depression Rating Scale, Geriatric Anxiety Scale and COMPASS-31. The method used to calculate change scores must be stated consistently for all outcomes.

