## Supplementary material for "WALK for Freezing of Gait in Parkinson’s Disease: A Sham-Controlled Home Rehabilitation Trial": participants data

Baseline demographic variables, disease characteristics, gait and balance outcomes (mZ-FOG total, Mini‑BEST, Functional Gait Assessment [FGA], Tinetti), and non‑motor symptom scores (HDRS, GAS, COMPASS) of patients with Parkinson’s disease enrolled in the intervention and sham group.

| **Name of the Patient** | **Group** | **Sex** | **Age** | **Disease Duration** | **H&Y** | **mZ-FOG total score** | **Mini-BEST** | **FGA** | **Tinetti** | **HDRS** | **GAS** | **COMPASS** |
| --- | --- | --- | --- | --- | --- | --- | --- | --- | --- | --- | --- | --- |
| 130003 | Intervention | Male | 64 | 2 | 3 | 6 | 18 | 18 | 19 | 6 | 11 | 7 |
| 130005 | Intervention | Male | 62 | 13 | 3 | 12 | 18 | 10 | 20 | 1 | 2 | 9 |
| 180002 | Intervention | Male | 70 | 30 | 2.5 | 13 | 17 | 16 | 14 | 3 | 11 | 12 |
| 220006 | Intervention | Female | 73 | 7 | 3 | 16 | 17 | 26 | 24 | 4 | 5 | 30 |
| 230004 | Intervention | Male | 59 | 13 | 4 | 29 | 20 | 13 | 23 | 5 | 7 | 11 |
| 240008 | Intervention | Male | 73 | 12 | 3 | 24 | 17 | 11 | 16 | 9 | 35 | 18 |
| 250001 | Intervention | Male | 67 | 17 | 3 | 24 | 11 | 13 | 12 | 20 | 29 | 13 |
| 330006 | Intervention | Male | 76 | 1 | 4 | 12 | 23 | 22 | 27 | 16 | 35 | 30 |
| 370000 | Intervention | Male | 67 | 10 | 3 | 22 | 14 | 19 | 18 | 7 | 16 | 13 |
| 370006 | Intervention | Male | 69 | 18 | 3 | 6 | 20 | 22 | 19 | 6 | 15 | 7 |
| 480002 | Intervention | Male | 66 | 1 | 3 | 16 | 17 | 12 | 19 | 6 | 12 | 10 |
| 530001 | Intervention | Male | 58 | 13 | 2 | 20 | 23 | 7 | 21 | 9 | 27 | 16 |
| 540005 | Intervention | Male | 75 | 11 | 3 | 16 | 17 | 6 | 26 | 7 | 9 | 17 |
| 550009 | Intervention | Male | 69 | 0.5 | 3 | 19 | 19 | 11 | 21 | 0 | 9 | 0 |
| 590000 | Intervention | Male | 66 | 3 | 3 | 7 | 26 | 24 | 23 | 9 | 26 | 16 |
| 630007 | Intervention | Male | 62 | 7 | 2.5 | 13 | 22 | 23 | 22 | 2 | 8 | 4 |
| 640009 | Intervention | Male | 64 | 21 | 2.5 | 9 | 14 | 17 | 22 | 8 | 9 | 8 |
| 650002 | Intervention | Male | 77 | 1 | 3 | 6 | 22 | 19 | 25 | 6 | 17 | 17 |
| 690009 | Intervention | Female | 63 | 4 | 3 | 13 | 19 | 10 | 19 | 7 | 14 | 0 |
| 740002 | Intervention | Male | 63 | 3 | 2 | 23 | 21 | 19 | 18 | 12 | 24 | 25 |
| 740005 | Intervention | Male | 65 | 15 | 2.5 | 29 | 18 | 11 | 16 | 7 | 14 | 11 |
| 930002 | Intervention | Male | 87 | 15 | 4 | 27 | 10 | 7 | 16 | 5 | 14 | 8 |
| 960001 | Intervention | Male | 71 | 16 | 4 | 25 | 25 | 15 | 22 | 10 | 24 | 13 |
| 990007 | Intervention | Male | 65 | 13 | 4 | 7 | 19 | 16 | 21 | 12 | 35 | 15 |
| 230003 | Sham | Female | 65 | 12 | 3 | 13 | 18 | 19 | 21 | 1 | 14 | 7 |
| 260002 | Sham | Male | 75 | 10 | 3 | 5 | 16 | 17 | 25 | 10 | 16 | 12 |
| 290001 | Sham | Male | 73 | 15 | 3 | 16 | 22 | 23 | 22 | 11 | 16 | 22 |
| 310004 | Sham | Male | 69 | 16 | 4 | 12 | 11 | 3 | 17 | 10 | 9 | 4 |
| 470006 | Sham | Female | 55 | 16 | 3 | 17 | 25 | 19 | 17 | 19 | 59 | 6 |
| 520007 | Sham | Male | 81 | 1 | 4 | 20 | 18 | 8 | 18 | 3 | 9 | 5 |
| 540004 | Sham | Male | 79 | 1 | 3 | 15 | 9 | 8 | 13 | 3 | 9 | 6 |
| 590008 | Sham | Male | 68 | 16 | 3 | 29 | 16 | 11 | 14 | 7 | 14 | 12 |
| 670003 | Sham | Male | 60 | 10 | 3 | 25 | 20 | 19 | 22 | 24 | 13 | 3 |
| 700000 | Sham | Male | 74 | 6 | 3 | 11 | 11 | 12 | 17 | 14 | 44 | 22 |
| 760009 | Sham | Male | 53 | 10 | 4 | 33 | 24 | 18 | 27 | 6 | 15 | 4 |
| 820006 | Sham | Male | 70 | 15 | 3 | 17 | 16 | 11 | 17 | 1 | 5 | 9 |
| 910007 | Sham | Male | 56 | 11 | 3 | 10 | 23 | 16 | 26 | 4 | 10 | 3 |
| 920001 | Sham | Female | 62 | 6 | 3 | 9 | 18 | 17 | 26 | 3 | 8 | 6 |
| 970002 | Sham | Female | 65 | 4 | 3 | 8 | 18 | 25 | 21 | 12 | 29 | 3 |
| 970005 | Sham | Female | 56 | 8 | 3 | 25 | 7 | 8 | 10 | 14 | 7 | 16 |
