## Supplementary material for "WALK for Freezing of Gait in Parkinson’s Disease: A Sham-Controlled Home Rehabilitation Trial": consort checklist

**The CONSORT reporting checklist**

*For checking that reports of randomised trials can be understood and used by everyone*

| **Item description** | **Location (or reason for not reporting)** |
| --- | --- |
| **Title and Abstract** | |
| 1a. Title Identification as a randomised trial. | Title, p.1 |
| 1b. Structured Abstract Structured summary of the trial design, methods, results, and conclusions. | Abstract, pp.1-2 |
| **Open Science** | |
| 2. Trial Registration Name of trial registry, identifying number (with URL) and date of registration. | Abstract; Methods, Study design and ethics, pp.1,3 |
| 3. Protocol and statistical analysis plan Where the trial protocol and statistical analysis plan can be accessed. | Data availability / trial registration statement, p.11 |
| 4. Data sharing Where and how the individual de-identified participant data (including data dictionary), statistical code and any other materials can be accessed. | Data availability, p.11 |
| **Funding and Conflicts of Interest** | |
| 5a. Funding Sources of funding and other support (eg, supply of drugs), and role of funders in the design, conduct, analysis, and reporting of the trial. | Funding, p.11 |
| 5b. Conflicts of interest Financial and other conflicts of interest of the manuscript authors. | Conflict of interest; patents, p.11 |
| **Introduction** | |
| 6. Background and rationale Scientific background and rationale. | Introduction, pp.2-3 |
| 7. Objectives Specific objectives related to benefits and harms. | Introduction, final paragraph, p.3 |
| **Methods** | |
| 8. Patient and public involvement Details of patient or public involvement in the design, conduct and reporting of the trial. | Not reported; add statement if no involvement |
| 9. Trial Design Description of trial design including type of trial (eg, parallel group, crossover), allocation ratio, and framework (eg, superiority, equivalence, non-inferiority, exploratory). | Methods, Study design and ethics; Randomisation, pp.3-4 |
| 10. Changes to trial protocol Important changes to the trial after it commenced including any outcomes or analyses that were not pre-specified, with reason. | Methods, Statistical analysis, p.5 |
| 11. Trial Setting Settings (eg, community, hospital) and locations (eg, countries, sites) where the trial was conducted. | Methods, Study design and ethics; Home-based programme, pp.3-5 |
| 12a. Participants Eligibility criteria for participants. | Methods, Participants, p.3 |
| 12b. Other If applicable, eligibility criteria for sites and for individuals delivering the interventions (eg, surgeons, physiotherapists). | Methods, Study design and home-based programme, pp.3-5 |
| 13. Intervention and comparator Intervention and comparator with sufficient details to allow replication. If relevant, where additional materials describing the intervention and comparator (eg, intervention manual) can be accessed. | Methods, WALK intervention and home-based programme, pp.4-5; Figure 1; Supplementary Document S2 |
| 14. Outcomes Prespecified primary and secondary outcomes, including the specific measurement variable, analysis metric, method of aggregation, and time point for each outcome. | Methods, Assessment schedule and outcomes, pp.4-5 |
| 15. Harms How harms were defined and assessed (eg, systematically, non-systematically). | Methods, Assessment scoring and safety, p.5 |
| 16a. How sample size was determined How sample size was determined, including all assumptions supporting the sample size calculation. | Methods, Statistical analysis, p.5 |
| 16b. Interim analyses and stopping criteria Explanation of any interim analyses and stopping guidelines. | Methods, Statistical analysis, p.5 |
| 17a. Sequence Generation Who generated the random allocation sequence and the method used. | Methods, Randomisation, concealment and masking, p.4 |
| 17b. Type of Randomisation Type of randomisation and details of any restriction (eg, stratification, blocking, and block size). | Methods, Randomisation, concealment and masking, p.4 |
| 18. Allocation concealment mechanism Mechanism used to implement the random allocation sequence, describing any steps to conceal the sequence until interventions were assigned. | Methods, Randomisation, concealment and masking, p.4 |
| 19. Implementation Whether the personnel who enrolled and those who assigned participants to the interventions had access to the random allocation sequence. | Methods, Randomisation, concealment and masking, p.4 |
| 20a. Who was blinded Who was blinded after assignment to interventions (eg, participants, care providers, outcome assessors, data analysts). | Methods, Randomisation, concealment and masking, p.4 |
| 20b. How blinding was achieved If blinded, how blinding was achieved and description of the similarity of interventions. | Methods, Randomisation, concealment and masking, p.4; Strengths and limitations, p.10 |
| 21a. Comparing groups Statistical methods used to compare groups for primary and secondary outcomes, including harms. | Methods, Statistical analysis, p.5 |
| 21b. Definition of who is included in each analysis Definition of who is included in each analysis, and in which group. | Methods, Statistical analysis; Results, Participants, pp.5-6; Figure 3 |
| 21c. Missing Data How missing data were handled in the analysis. | Methods, Statistical analysis, p.5 |
| 21d. Additional Analyses Methods for any additional analyses, distinguishing pre-specified from post hoc. | Methods, Statistical analysis, p.5; Supplementary Tables S2-S4 |
| **Results** | |
| 22a. Participant Numbers For each group, the numbers of participants who were randomly assigned, received intended intervention, and were analysed for the primary outcome. | Results, Participants and baseline characteristics, p.6; Figure 3 |
| 22b. Losses and exclusions For each group, losses and exclusions after randomisation, together with reasons. | Results, Participants and baseline characteristics, p.6; Figure 3 |
| 23a. Dates Dates defining the periods of recruitment and follow-up for outcomes of benefits and harms. | Methods, Study design and ethics, p.3 |
| 23b. Reasons for stopping recruitment If relevant, why the trial ended or was stopped. | Results, Participant flow / trial completion statement, p.6 |
| 24a. As Administered Intervention and comparator as they were actually administered. | Results, Feasibility and safety, p.6; Supplementary Document S2 |
| 24b. Concomitant Care Concomitant care received during the trial for each group. | Methods, Participants, p.3 |
| 25. Baseline Data A table showing baseline demographic and clinical characteristics for each group. | Table 1 |
| 26. Numbers analysed, outcomes, and estimation For each primary and secondary outcome, by group: number included in the analysis; number with available data; result for each group; estimated effect size and precision; and for binary outcomes both absolute and relative effect size. | Results, pp.6-7; Figure 4; Supplementary Tables S1-S5 |
| 27. Harms All harms or unintended events in each group. | Results, Feasibility and safety, p.6 |
| 28. Ancillary Analyses Any other analyses performed, including subgroup and sensitivity analyses, distinguishing pre-specified from post hoc. | Results, pp.6-7; Supplementary Figure S1; Supplementary Tables S2-S4 |
| **Discussion** | |
| 29. Interpretation Interpretation consistent with results, balancing benefits and harms, and considering other relevant evidence. | Discussion, pp.7-10 |
| 30. Limitations Trial limitations, addressing sources of potential bias, imprecision, generalisability, and, if relevant, multiplicity of analyses. | Discussion, Strengths and limitations, pp.10-11 |

*Note: Page references correspond to the current manuscript and should be refreshed after final pagination.*

**Checklist citation:** Hopewell S, Chan AW, Collins GS, et al. CONSORT 2025 statement: updated guideline for reporting randomised trials. BMJ. 2025;389:e081123.
