## Supplementary material for "WALK for Freezing of Gait in Parkinson’s Disease: A Sham-Controlled Home Rehabilitation Trial": S1

| ***Table S1 — Repeated-Measures ANOVA (Mixed Design)*** | | | | | | |
| --- | --- | --- | --- | --- | --- | --- |
| ***Part A: Descriptive Statistics (Mean ± SD)*** | | | | | | |
| **Group** | **n** | **S0** | **S8 without device** | **S8 with device** | **S16 without device** | **S16 with device** |
| **Sham** | 16 | 16.63 ± 8.37 | 12.38 ± 9.07 | 11.00 ± 6.48 | 14.00 ± 8.48 | 10.19 ± 8.19 |
| **Intervention** | 24 | 16.42 ± 7.64 | 11.79 ± 7.54 | 8.33 ± 6.35 | 10.88 ± 7.27 | 7.21 ± 6.47 |
| ***Part B: ANOVA Results (Greenhouse-Geisser corrected df)*** | | | | | | |
| **Effect** | **df (GG corrected)** | **F** | **p-value** | **Partial η²** | **Interpretation** | |
| **Condition** | 2.826, 107.40 | 15.94 | < .001 | 0.295 | *Significant within-subjects effect; sphericity violated (Mauchly p<.001), ε=0.706, GG corrected* | |
| **Group (between-subjects)** | 1, 38 | 0.904 | 0.348 | 0.023 | *No overall difference between groups across the five assessment conditions.* | |
| **Condition × Group (interaction)** | 2.826, 107.40 | 0.847 | 0.465 | 0.022 | *No significant group-by-time interaction (non-significant)* | |
| *Note: GG = Greenhouse-Geisser sphericity correction.*  The analysis included 40 participants with complete mZ-FOG scores across all five conditions: active WALK, n=24; sham WALK, n=16. Group was the between-participant factor and Condition the within-participant factor. The Group × Condition interaction was the main comparison. Greenhouse–Geisser correction was applied where the assumption of sphericity was not met. Partial *η²* is reported as the effect-size measure.  *Partial η² interpreted as small (0.01), medium (0.06), large (0.14).* | | | | | | |
