## Supplementary material for "WALK for Freezing of Gait in Parkinson’s Disease: A Sham-Controlled Home Rehabilitation Trial": S2

| ***Table S2 — Linear Mixed Model Results*** | | | | | | |
| --- | --- | --- | --- | --- | --- | --- |
| ***Part A: Fixed Effects*** | | | | | | |
| **Effect** | **Estimate** | **SE** | **95% CI Lower** | **95% CI Upper** | **t-value** | **p-value** |
| **Intercept** | 16.625 | 1.881 | 12.938 | 20.312 | 8.838 | < 0.001 |
| **Group: Intervention** | -0.208 | 2.429 | -4.968 | 4.552 | -0.086 | 0.932 |
| **Condition (S8 without device)** | -4.25 | 1.667 | -7.512 | -0.982 | -2.549 | 0.012 |
| **Condition (S8 with device)** | -5.625 | 1.667 | -8.892 | -2.357 | -3.374 | < 0.001 |
| **Condition (S16 without device)** | -2.625 | 1.667 | -5.892 | 0.643 | -1.575 | 0.117 |
| **Condition (S16 with device)** | -6.437 | 1.667 | -9.705 | -3.169 | -3.861 | < 0.001 |
| **Group × S8 (without device)** | -0.375 | 2.152 | -4.593 | 3.843 | -0.174 | 0.862 |
| **Group × S8 (with device)** | -2.458 | 2.152 | -6.676 | 1.758 | -1.142 | 0.255 |
| **Group × S16 (without device)** | -2.917 | 2.152 | -7.135 | 1.302 | -1.355 | 0.177 |
| **Group × S16 (with device)** | -2.771 | 2.152 | -6.989 | 1.447 | -1.287 | 0.2 |
| ***Part B: Variance Components (Random Effects)*** | | |  |  |  |  |
| **Component** | **Variance** | **SD** |  |  |  |  |
| Subject (random intercept) | 34.385 | 5.864 |  |  |  |  |
| Residual | 22.234 | 4.715 |  |  |  |  |
| *Note:* ***Sham WALK and baseline without the device were the reference categories. Fixed-effect estimates are expressed in mZ-FOG points; negative estimates indicate lower freezing scores relative to the reference category. The model included a participant-level random intercept.***  *Model fitted via restricted maximum likelihood (REML). Reference level: Sham group at Baseline (S0). Sig. codes: *** p<0.001, ** p<0.01, * p<0.05, † p<0.10* | | | | | | |
