## Supplementary material for "WALK for Freezing of Gait in Parkinson’s Disease: A Sham-Controlled Home Rehabilitation Trial": S3

| ***Table S3 — Robustness Analyses: Bootstrapped Contrasts & Rank-Normal Models*** | | | | | | | |
| --- | --- | --- | --- | --- | --- | --- | --- |
| ***Bootstrap (BCa, n=2000 resamples) and rank-normal pairwise contrasts. Within-group: with device minus without device. Between-group: Intervention minus Sham.*** | | | | | | | |
| **Comparison** | **Group** | **Bootstrap Mean Δ** | **BCa 95% CI Lower** | **BCa 95% CI Upper** | **Bootstrap p value** | **Rank-normal Estimate**  **(Hedges g)** | **Interpretation** |
| S8 with device − S8 without device (within-session change at S8) | **Intervention** | -3.458 | -5 | -2.5 | < 0.001 | -1.044 | *Significant Improvement* |
| S8 with device − S8 without device (within-session change atS8) | **Sham** | -1.375 | -4.75 | 1.307 | 0.383 | -0.196 | *Non-significant* |
| S16 with device − S16 without device (within-session change at  S16) | **Intervention** | -3.667 | -5.708 | -1.875 | < 0.001 | -0.73 | *Significant Improvement* |
| S16 with device − S16 without device (within-session change at  S16) | **Sham** | -3.812 | -5.757 | -2.125 | < 0.001 | -0.896 | *Significant Improvement* |
| S8 with device − S0 (change to S8) | **Intervention** | -8.083 | -10.875 | -5.625 | <0.001 | -1.167 | *Significant Improvement* |
| S8 with device − S0 (change to S8) | **Sham** | -5.625 | -8.75 | -2.625 | <0.001 | -0.816 | *Significant Improvement* |
| S16 with device − S0 (overall change) | **Intervention** | -9.208 | -11.958 | -6.375 | < 0.001 | -1.278 | *Significant Improvement* |
| S16 with device − S0 (overall change) | **Sham** | -6.438 | -9.875 | -3.375 | < 0.001 | -0.87 | *Significant Improvement* |
| Intervention − Sham at S16 with device | **Between groups** | -2.979 | -7.915 | 1.312 | 0.219 | -0.406 | *Non-significant* |
| *Note:* *Within-session changes compare the score with the assigned device against the score without the device at the same assessment. Negative values indicate lower mZ-FOG scores in the later, with-device condition. Between-group contrasts are calculated as active WALK minus sham WALK; negative values favour active WALK because lower scores indicate less severe freezing. Bootstrap confidence intervals were calculated using 2,000 resamples.*  *BCa = bias-corrected and accelerated bootstrap. Rank-normal model uses rank-transformed mZ-FOG total scores with Bonferroni-adjusted p-values. Hedges g: small 0.2, medium 0.5, large 0.8.* | | | | | | | |
