## Supplementary material for "WALK for Freezing of Gait in Parkinson’s Disease: A Sham-Controlled Home Rehabilitation Trial": S4

| ***Table S4 — ANCOVA: Baseline-Adjusted Group Differences*** | | | | | | | | |
| --- | --- | --- | --- | --- | --- | --- | --- | --- |
| ***OLS ANCOVA: Outcome ~ Group + Baseline (S0). Adjusted means estimated at grand-mean S0.*** | | | | | | | | |
| **Outcome (Timepoint)** | **Adj. Mean (Active WALK)** | **Adj. Mean (Sham WALK)** | **Adjusted Diff**  **(Active WALK − Sham WALK)** | **95% CI Lower** | **95% CI Upper** | **p-value** | **Effect Size**  **(Cohen's d)** | **Interpretation** |
| **S8 without device** | 11.84 | 12.3 | -0.46 | -4.87 | 3.96 | 0.834 | -0.068 | *Non-significant* |
| **S8 with device** | 8.37 | 10.94 | -2.57 | -5.99 | 0.86 | 0.137 | -0.49 | *Non-significant* |
| **S16 without device** | 10.93 | 13.91 | -2.98 | -6.63 | 0.66 | 0.106 | -0.535 | *Non-significant* |
| **S16 with device** | 7.25 | 10.12 | -2.87 | -6.76 | 1.02 | 0.144 | -0.482 | *Non-significant* |
| *Note: Values are estimated means adjusted for baseline mZ-FOG score. Between-group differences are calculated as active WALK minus sham WALK. Negative values favour active WALK because lower mZ-FOG scores indicate less severe freezing. Standardised differences follow the same direction.*  *Positive difference = Active WALK scored higher. Cohen's d computed from adjusted difference / pooled SD. Covariate: Baseline mZ-FOG total score (S0).* | | | | | | | | |
