## Supplementary material for "WALK for Freezing of Gait in Parkinson’s Disease: A Sham-Controlled Home Rehabilitation Trial": S5

| ***Table S5 — Secondary clinical outcomes: change from baseline at S8 and S16.*** | | | | | | | |
| --- | --- | --- | --- | --- | --- | --- | --- |
| ***Mean change (Δ) from Baseline ± 95% CI for each secondary outcome and timepoint.*** | | | | | | | |
| **Outcome** | **Group** | **Δ S8** | **95% CI** | **p-value** | **Δ S16** | **95% CI** | **p-value** |
| **Tinetti (Balance & Gait)** | | | | | | | |
| S0: 20.1 ± 3.72 | Intervention | -3.71 | [-7.07, -0.35] | 0.020* | -4.42 | [-7, -1.83] | < 0.001*** |
| S0: 19.6 ± 5.02 | Sham | -4.56 | [-8.68, -0.45] | 0.019* | -3.69 | [-6.86, -0.52] | 0.012* |
| **MiniBEST (Balance)** | | | | | | | |
| S0: 18.6 ± 3.93 | Intervention | -0.25 | [-2.87, 2.37] | 1 | -1.58 | [-4.89, 1.72] | 1 |
| S0: 17.0 ± 5.33 | Sham | -1.94 | [-5.14, 1.27] | 0.99 | -2.75 | [-6.8, 1.30] | 0.6 |
| **FGA (Functional Gait Assessment)** | | | | | | | |
| S0: 15.3 ± 5.71 | Intervention | -1.38 | [-4.88, 2.14] | 1 | -3.54 | [-7.53, 0.45] | 0.13 |
| S0: 14.6 ± 6.11 | Sham | -2.13 | [-6.42, 2.17] | 1 | -2.88 | [-7.77, 2.02] | 1 |
| **HDRS (Depression)** | | | | | | | |
| S0: 7.4 ± 4.50 | Intervention | 1.46 | [-1.78, 4.7] | 1 | 1.33 | [-2.05, 4.72] | 1 |
| S0: 8.9 ± 6.65 | Sham | 3.44 | [-0.53, 7.41] | 0.15 | 3.44 | [-0.705, 7.58] | 0.2 |
| **COMPASS (Autonomic Symptoms)** | | | | | | | |
| S0: 12.9 ± 7.73 | Intervention | 3.667 | [0.37, 6.97] | 0.019* | 4.125 | [1.05, 7.2] | 0.002** |
| S0: 8.8 ± 6.37 | Sham | 3.125 | [-0.91, 7.16] | 0.3 | 2.125 | [-1.64, 5.89] | 1 |
| **GAS (Anxiety)** | | | | | | | |
| S0: 17 ± 9.93 | Intervention | 3.375 | [-3.574, 10.32] | 1 | 3.167 | [-2.916, 9.25] | 1 |
| S0: 17.31 ± 14.7 | Sham | 4.75 | [-3.761, 13.26] | 1 | 2.813 | [-4.638, 10.26] | 1 |
| *Note: Δ = change from Baseline (S0; baseline minus follow-up). 95% CI from mixed model or t-test. Duplicate the shaded block for each additional secondary outcome. p-values Bonferroni-corrected for number of secondary outcomes if required.*  *Change was calculated as baseline minus follow-up. Positive values therefore indicate improvement for mZ-FOG and symptom scales, where lower follow-up scores are favourable. Negative values indicate improvement for Mini-BESTest, Functional Gait Assessment and Tinetti/POMA, where higher follow-up scores are favourable.* | | | | | | | |
