## Supplementary figures and images for "WALK for Freezing of Gait in Parkinson’s Disease: A Sham-Controlled Home Rehabilitation Trial"

### Supplement F1

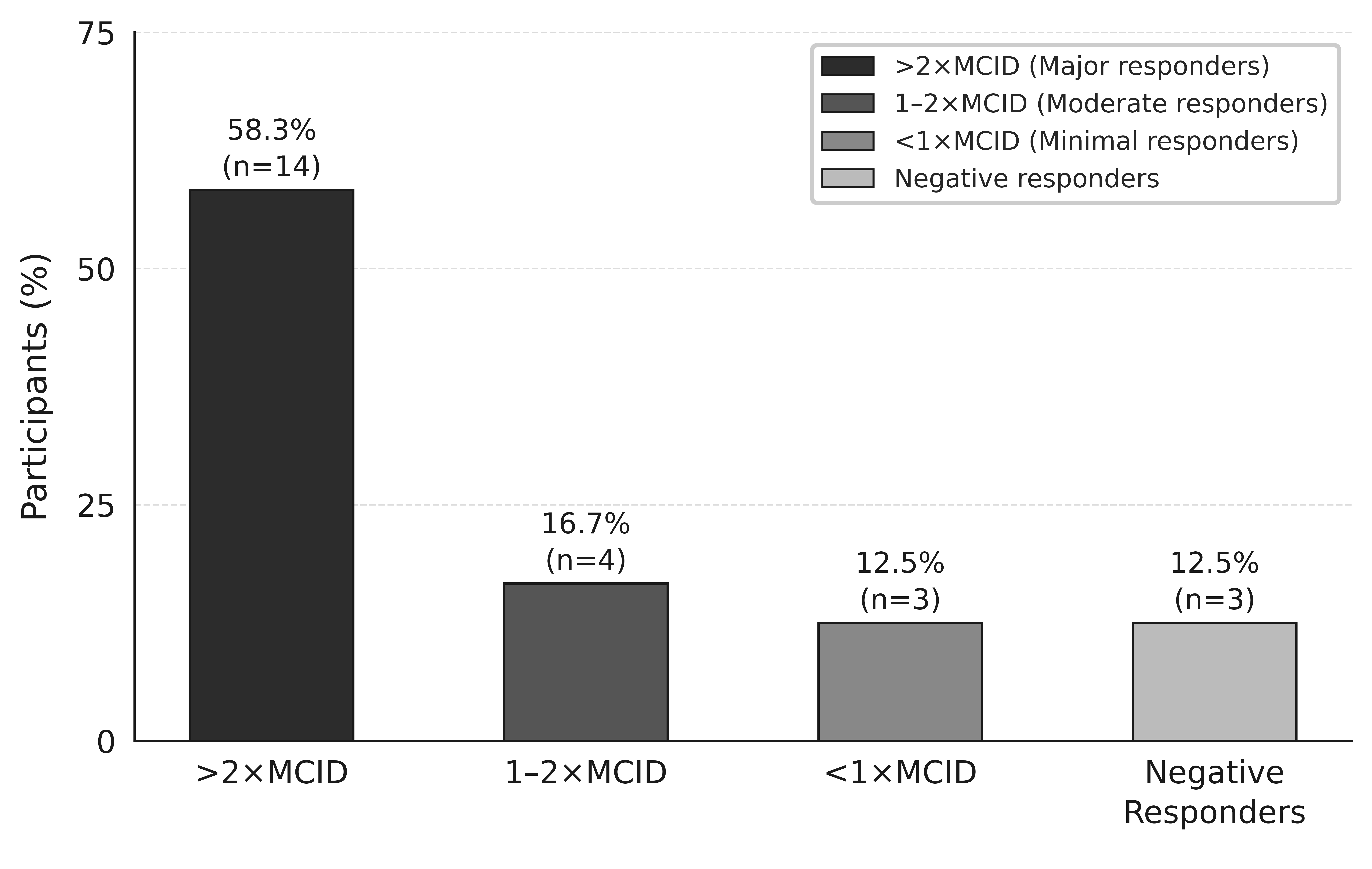
